# Clumpiness: Modeling the Impact of Social Dynamics on COVID-19 Spread

**DOI:** 10.1101/2021.04.12.21255149

**Authors:** Ben Goertzel, Cassio Pennachin, Deborah Duong, Matt Iklé, Michael Duncan, James Boyd, André Senna, Ramon Durães

## Abstract

We present an agent based simulation model configured for exploring the dynamics of disease spread in the context of agents that group together through homophily, the principle of “like attracts like”. To study the properties of this model, we introduce two novel social network inter-connectivity measures, “clumpiness” and “hoprank,” that are the same basic concept defined at global and local levels, respectively. The measures may be computed from samples of readily available demographic data, and are useful for measuring probabilistic packet transmission through social networks. In three studies we apply clumpiness to measure the effects, on COVID-19 transmission, caused by social networks of both homophilic physical proximity and homophilic information replication. The particular characteristic we are interested in about disease transmission is herd immunity, the percentage of a population that has to be immune in order to prevent infection from spreading to those who are not. Two studies demonstrate innovations measuring herd immunity levels and predicting future outbreak locations, procedures relevant to epidemiological control policy. In the first study, we look at how homophilic physical proximity networks form natural bubbles that act as frictive surfaces that affect the speed of transmission of packets and influence herd immunity levels. In the second study, we test clumpiness in homophilic proximity social networks as a predictor of future infection outbreaks at the level of individual schools, restaurants, and workplaces. Our third study demonstrates that protective social bubbles form naturally from homophilic information replication networks, and enhance the natural bubbles that come from the homophilic physical proximity networks. Accurate description of this information environment lays the foundation for epidemiological messaging policy formation.

## 1 Introduction

We present an agent based simulation model configured for exploring the dynamics of disease spread in the context of agents that group together through homophily, the principle of “like attracts like”. To study the properties of this model, we introduce two novel social network interconnectivity measures, “clumpiness” and “hoprank”, that are the same basic concept defined at global and local levels, respectively. While contact tracing uses contact networks to identify cases in an effort to interrupt disease progression, we use graphical networks to predict future transmission trajectories of individuals and groups of individuals. In the study most similar to ours, Block et al. [5] advocate using hop distance of shortest paths to relate social network structures to infection transmission. We build upon this state of the art approach with a probabilistic measurement of social connectivity that we call clumpiness. Our approach uses the most probable path rather than the shortest path, a modification that facilitates more accurate forecasting and more effective policy development.

With clumpiness, we attempt to precisely define a measure of population interaction heterogeneity. Mathematically, we calculate a network average of node proximities, by defining clumpiness as 1 - the network average of the reciprocal of the number of hops necessary to cross the most probable path between individuals in the network (see definition 1.1.) As a result, for any network, we have 0 ⩽ *clumpiness ⩽*1. At one extreme, corresponding to *clumpiness* = 1, is the case in which every individual is isolated from (i.e. has edge probability 0 to) every other individual. At the other extreme, corresponding to *clumpiness* = 0, everyone is connected, with edge probability 1, to everyone else. Such a measure necessarily relies not just on population size and the number of communities, but also on the size distribution of those communities, and on the distributions of edges and edge weights.

### Definition 1.1

Mathematically, if the number of nodes is *N*, then

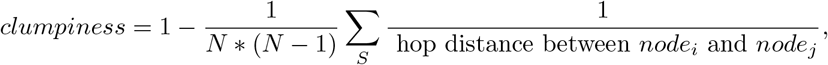

where the set *S* is the set of all pairs (*node*_*i*_, *node*_*j*_) of node pairs *i* ≠ *j* for which a path exists from *node*_*i*_ to *node*_*j*_.

For a given population size, a network with many small tightly-knit communities, connected loosely to each other will usually be more clumpy than a network consisting of a single large tightly-connected community. With a larger number of small communities, there will be more people outside one’s own community so that, on average, the number of hops separating individuals will also be larger (see figures 1 and 2).

**Figure 1:**
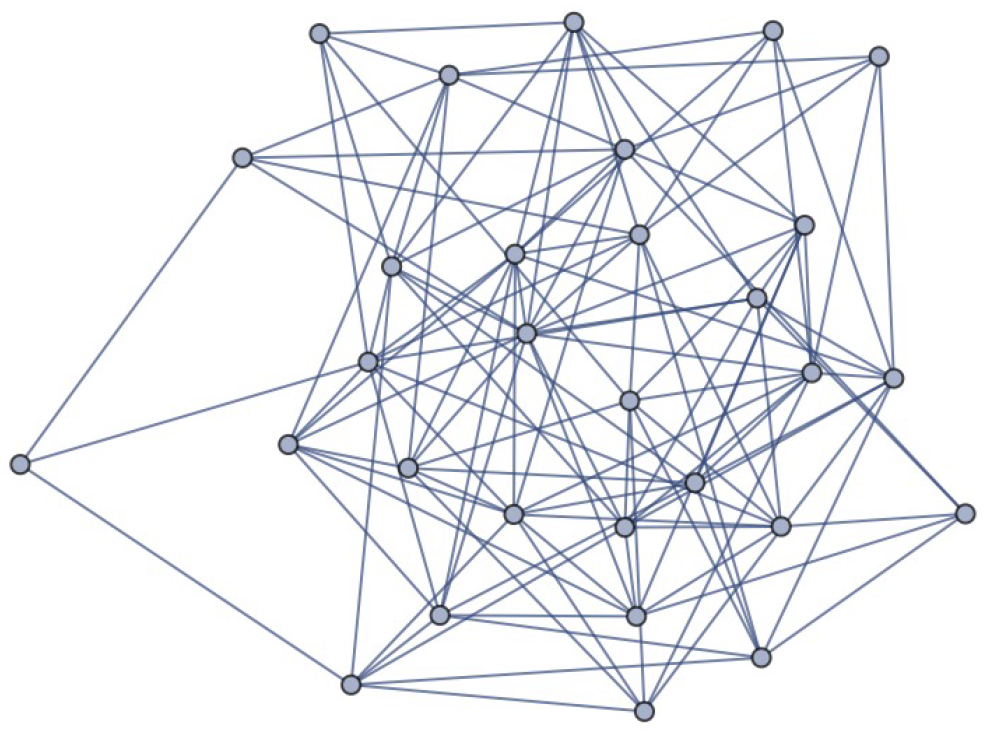
Low clumpiness with 30 nodes.

**Figure 2:**
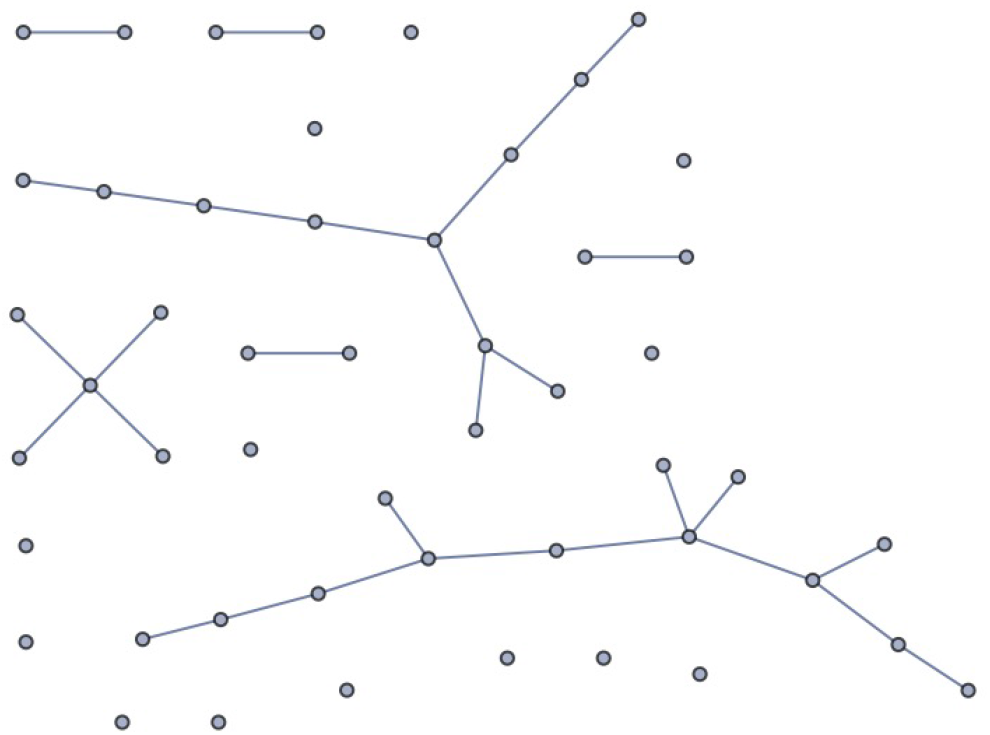
High clumpiness with 30 nodes.

Clumpiness is thus a network property that measures interconnectivity based on social proximity. It is useful in epidemiological studies of physical viral transmission as well as in sociological studies of memetic transmission. Clumpiness has some characteristics of hop-distance measures such as “average shortest path” and some characteristics of probabilistic measures like pagerank. Specifically, we can apply a probabilistic component to a distance based temporal component to predict when packets are likely to arrive. Pagerank measures the total probability of all possible paths but can not include a distance based temporal component because it would be intractable to include path lengths in the eigenvector calculation. Clumpiness makes the tradeoff, for tractability, of including only the most probable path between two nodes. It nonetheless yields good estimates of the arrival times of packets.

As a measure of interconnectivity of nodes with dynamic connections, inclusion of temporary disconnections is important to our clumpiness definition. Including an individual’s disconnections and connections, both probable and improbable, improves the measurement accuracy of an individual’s isolation.

A social proximity network connects persons to locations (physical or virtual) that may be shared with other persons, weighted with the probability of transmission of packets (e. g. viral or memetic) between persons at the same location. Individuals frequenting specific locations help create a network that is useful for prediction. To efficiently compute the maximally probable path we use Dijkstra’s shortest path algorithm [9], applied to the negative logarithm of the individual probabilities of transmission between agents. We consider persons to be co-located if they pass through a common location during the same day, though possibly at different times, by factoring any lack of simultaneity into the probability weights. Such a flexible definition of co-location facilitates the modeling of fomites or aerosols for viral transmission, enables easier data collection, and allows the representation of potential transmission.

Clumpiness can be used to measure the drag the network puts on transmissions since it measures the number of hops, and hence amount of time, packets require to get from one person to another. A virus takes time to incubate, for example, or memes may take time to be internalized by agents before they spread it to others. In the case of viruses this drag affects the herd immunity level of the society, the percentage of a population that has to be immune in order to prevent infection from spreading to those who are not.

One key social force that we posit as causing social networks to be clumpy is homophily; that is, people tend to stay in proximity to people most socially similar to themselves ^1^. We purposely leave these features as abstract but they may be construed as any social marker that others recognize, including race, gender, generation, ethnicity, education level, wealth, religion, occupation, etc. We use cosine similarity of feature vectors of these traits as the basis for computing the level of homophily in the network.

Homophily is important in our measurements of infection as it can be used to infer clumpiness. If sufficient data on the locations where individuals spend time is not available, then information about types of locations where types of individuals go can substitute. Clumpiness can be measured in real world data available on social media platforms that profile persons and the locations that they frequent. Homophily can be measured with the same data or, because it is correlated with clumpiness, can be used to impute missing data. We use homophily in our simulation studies to generate clumpiness.

We seek to describe a preexisting frictive social surface along which packets travel. While we can examine clumpiness as a network trait calculated via measurements between individuals of the network, we can also characterize the contribution of individuals or groups to the clumpiness of their network in a measurement of their own isolation from other individuals and groups in their network. We thus distinguish two levels of social interconnectivity: a global network level and a local mutually exclusive sub-network level. The clumpiness of a network is the average hoprank of its individuals. Hoprank is similar to closeness centrality, with the most probable path replacing the shortest path in measuring “closeness”. This change makes hoprank applicable regardless of whether or not packets travel the shortest distance. Infection is one thing that doesn’t necessarily travel on the shortest path, making most probable path a better predictor.

If those hopranked subsets are individuals, we rank the time of packet arrival to individual persons or locations. If the packets along the network are viruses, then hoprank predicts the general order in which agents are likely to be exposed to the virus. Hoprank itself doesn’t include the delay times for either virus incubation or message interpretation, but can be multiplied by these for an ETA. Like clumpiness, hoprank values range from zero to one. Agents with lower hopranks are likely to be exposed sooner to the disease than are agents having higher hopranks. Such information can be useful for prioritizing vaccinations. We will show that hoprank is particularly effective when knowledge of infected communities is taken into account. In such cases, individuals are hopranked according to their distance from communities known to be infected.

## 2 Previous Studies on the Impact of Social Structure on Infection

### 2.1 Transmission and Modeling Population Heterogeneity

The classical approach to modeling epidemics uses compartment models, with individuals passing between “compartments” of susceptible, infectious, and recovered/immune (SIR) partitions of the population, usually analyzed as a system of differential equations. This sort of model can be expanded to account for exposed but not yet infectious people (SEIR) or time limited immunity (SIRS), with the order of letters corresponding to the flow of people through the compartments. More recent work adds spatially and socially separated sub-populations with variable levels of within group and between group mixing. For example, Appolloni, et al. [4], analyze the early spread of H1N1 influenza virus from Mexico to Europe in the 2009 pandemic using an age partition of youth and adult groups. This subject stratification permits the model to account for the increased tendency of children to mix preferentially with other children (homophily), while adults on average mix with both children and adults (assortative versus proportional mixing,) children having less preexisting immunity on average than adults, and adults traveling between spatially separate regions more often than children.

Graph theory permits the modeling of epidemics with smaller compartments allowing for the capture of greater population heterogeneity in the level of transmission events between individuals. Stegehuis, et al [20], show that community structures defined as clusters in real-world networks can accelerate or inhibit spread among the network nodes by comparing percolation on random graphs that preserve the edge density distribution defining the empirical community clusters. They further conclude that inter-cluster edge density dominates internal community structure in determining global percolation behavior. Loyal and Chen [16] use statistical network analysis to demonstrate the effects of population heterogeneity in the context of a standard SEIR compartment model applied to contact networks. Using real contact data from a primary school they model the potential decrease in disease transmission achieved by a simulated intervention that divides the students into two groups attending school on alternate days.

The ever-increasing availability of computing power provides an alternative to traditional graph theoretical and statistical approaches: network diffusion analysis via agent based modeling. Interactions among large populations with individuals distinguished by many dimensions of heterogeneity can be simulated in realistic environments to model complex nonlinear dynamics and predict the effects of potential intervention strategies that are not tractable analytically. Block, et al [5], use an agent based simulation to model strategies to limit social interactions short of complete household isolation, thus potentially mitigating the psychological and economic impacts of complete lockdown while preventing health care capacity from becoming overwhelmed.

### 2.2 Empirical measures of social mixing

Read, et al. [19] provide a good overview of the problem of measuring population social interaction dynamics. Estimates of model parameters governing patterns of communicable disease transmission have traditionally been based on social contact surveys, in which members of a sample population from the area under study are queried about their contact history during a specific time period. Low-power radio proximity sensors have also been employed, though this is only feasible in limited environments such as schools and other institutional settings. More recently, in response to the Covid 19 epidemic, Google and Apple have made APIs available for myriad bluetooth based contact tracing apps [1]. Secondary data analysis combines such individual-level measurements with socio-demographic data to infer population level estimates for epidemic modeling. A recent literature review by Hoang, et al. [12], found consistent results across 65 studies, though most surveys were done in high-income countries. Age strata, household size, and time scheduling (such as day of the week) were identified as major determinants of contact patterns. They note that a major source of study variability is discrepancy across studies in the definition of contact. This includes differences in the minimal distance and modality (such as proximity or physical contact that define interaction.) These details are key to relating contacts to potential for spread of specific infectious agents; such as contact via droplet or aerosol transmission.

A notable investigation by Strömgren, et al, [21] develops a four category typology of mixing sites from a cross-sectional contact diary survey of the Swedish general population by clustering contact events by frequency of occurrence and average duration. They observe the most frequently occupied site with the longest duration and highest chance of physical contact to be the family home. The home, social network hubs (gathering sites for family and friends) and fixed activity sites such as schools, work places, and sports venues had the longest average duration of contact as well as high likelihood of physical contact, while fixed activity sites and “trading plazas,” which included public transport, restaurants and stores, had the greatest average number of contacts. One of the largest studies to date is POLYMOD [17], which analyzes a representative sample of 7,290 people from eight European countries who recorded a total of 97,904 contacts over a 24 hour period, specifying location and duration of contact, and the age and sex of contact parties. This study offers a major validation of age assortativity as a key structure in population mixing patterns. Prem, et al. [18], expand the results to 152 countries by predicting age group by location contact matrices using parameters from a Bayesian hierarchical model of the POLYMOD results combined with other comprehensive regional study results and data on household structure, age composition, labor force participation, and school enrollment made available by other countries and used for extrapolative purposes. Large cross-sectional contact surveys are still relatively rare, especially for low income countries, due to the significant resource requirements.

## 3 Agent-Based COVID-19 Model Description

To improve upon our current understanding of pandemic spread we have integrated both demographic stratification and fundamental social interaction dynamics within our COVID-19 agent-based simulations, allowing us to fully test our social network measures of clumpiness and hoprank in a simulated population of agents. It is the interaction dynamics, determined by agent choices, that leads to the particular course of pandemic spread across the network. While in our current implementation, agent decisions are probabilistically determined based on a set of particular parameter values, planned future implementations would allow for truly intelligent reinforcement learning (RL) agents.

Few other agent based models adopt social structural and behavioral principles from the social sciences as their focus, although some share the detailed demographic stratification. The June model [6] uses detailed demographic and survey data of the population of England, coupled with Bayesian emulation, to calibrate a statistically similar population at a granular level. The Covasim simulation [13] uses social network implementation of stratifications for efficient computation. Other models have a few social features such as accounting for whether a contact is in the household [7, 15, 14]; accounting for the age and clustering of contacts within the household [2, 7, 14]; and accounting for use of schools and workplaces based on census and time-use data [2]. However, none use social behavioral patterns such as social “herding” (the propensity to copy one’s friends), risk tolerance or homophily, as causal processes.

Our model also allows tractable exploration and predictive modeling of the differential impact of various policy decisions. The underlying simulations we report here can be overlaid on maps of specific cities, states or nations. Coupled with region-specific data streams that include demographic, economic, business, governmental, educational, medical, transportation, and/or other infrastructure information, our simulations can illustrate, in detail, the evolution of COVID-19 dynamics within particular regions. Please see appendix 1 and 2 for additional details of our simulation setup.

## 4 Clumpiness Simulations and Results

### 4.1 Setup

In our simulation studies we generate various levels of clumpiness to test against infection by creating relationships at corresponding levels of homophily. Different societies have different amounts of population intermixing, and to cover them we have modeled a continuum of homophily scenarios. We control the level of homophily in the social networks of a simulation scenario with two tools: scikit learn’s make blobs function to create blobs of feature vectors representing communities of similar agents, and a choice function that assigns agents to social relationships such as family, worker, school, and friend relationships. We then relate homophily to clumpiness and clumpiness to infection outcomes.

### 4.2 The Choice Function

We used a choice function similar to the one used in Duong and Reilly [10] to determine the amount of homophily between agents that frequent the same locations. Our choice function takes”temperature” parameter values ranging from −1 (the frozen state) to 1 (the random state). A temperature of −1 dictates that agents can only be chosen from an agent pool for collocation with other agents that have an absolute maximal level of feature vector cosine similarity. A temperature of 0 means choosing proportionally to feature vector cosine similarity (also known as roulette wheel choice). A temperature of 1 denotes a random uniform sample from the pool. Between temperatures 0 and 1, choices are determined by a weighted-by-temperature average of uniform and roulette wheel distributions. For temperatures T between −1 and 0, inclusively, choices are determined via a random function for which the number of possible outcomes of the random variable X(T) varies upper semi-continuously, with the set of possible outcomes at temperature T including only elements accounting for the uppermost 1+T probabilities.

Specifically, we first order the elements in decreasing order of their probabilities, and include elements one-by-one until their accumulated probabilities first reach 1+T, truncating any probability value of the last added element added in excess of 1+T. We next renormalize these values for the chosen elements and use the resultant probabilities in roulette wheel selection.

As an example, consider the case with three elements in which the elements have probabilities 0.5, 0.3, and 0.2, and in which T = − 0.4, and hence 1 + T = 0.6. The elements one through three are already listed in decreasing order of their probabilities. We include only elements one and two, with portions of their probabilities corresponding to 0.5 and 0.1 respectively. Upon renormalizing, the probability of element 1 becomes 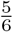 and that of element 2 becomes 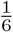. We then use roulette selection to choose among those elements, proportionate to those two probabilities.

In our simulation, we use scikit-learn’s make blobs function to assign a social marker feature vector of length *n*_*features*_ to each location (including homes, schools, offices, restaurants, etc.) and to each agent. We use the choice function to group families of related agents as well as to assign agents with feature vectors to locations. The blobs, as groups of feature vectors that are similar, represent similar agents. Near the frozen state, all agents are maximally close to the vector of the room and thus maximally close to each other. Temperature is an independent variable in our scenarios, creating homes, schools, offices and restaurants of various levels of homophily and as a result, various levels of clumpiness.

## 5 Clumpiness Simulation Experiments

We conducted experiments to study three separate but related phenomena. In Study 1, we primarily explore the relationship between homophily and infection outcomes, paying particular attention to tipping points in which small changes in social behaviors can result in large changes in infection, so that policy efforts may be more informed and better targeted. In our second study we explore whether hoprank could provide better predictions than the current state-of-the-art pagerank algorithm of the order in which the disease will arrive at particular schools, workplaces, and restaurants. Such information could be useful in prioritizing groups for vaccination campaigns. Finally, in our third study, we examine the effects of individual herding and risk tolerance behaviors on herd immunity and infection levels.

### 5.1 Study 1: Homophily and Infection

To examine the relationship between homophily and infection rates, we first generate scenarios at different homophily levels. We use 1000 agents, grouped into 20 communities by similarity in each run. We have 80 home districts forming 4 school districts. We use 40 temperature values and conduct 20 runs at each temperature to assure statistical significance.

Figure 3 displays the exponential relationship between temperature and homophily, with homophily measured via cosine vector similarity. The figure suggests that opening up social structure to allow more choice results in more intermixing. Each dot represents the average of 20 runs at the same temperature. Intuitively, homophily decreases exponentially as temperature rises because each new choice matters more when there are fewer choices than when there are already many choices. Here a choice is made when agents visit the same classroom, office, home, or restaurant.

**Figure 3:**
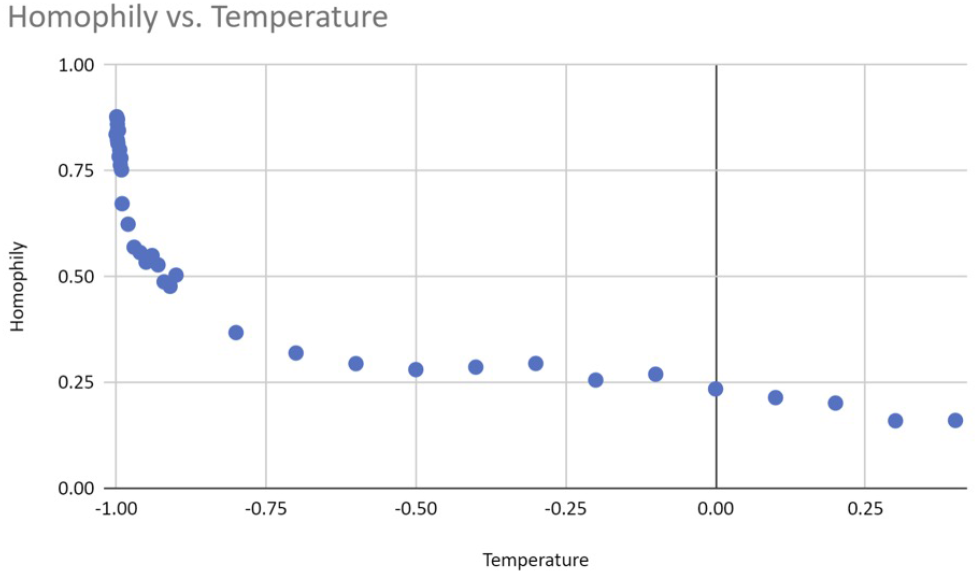
The relationship between temperature and homophily.

Our simulations show that both the peak of infection and the herd immunity level decrease as homophily increases. Figure 4 displays an example graphical output, including 95% confidence intervals, of a series of repeated runs with identical parameter settings. We also estimate that herd immunity occurs at roughly 20% of the population, as this is the height of the inflection point of the cumulative recovered (green) curve. Figures 5 and 6 illustrate that homophily levels in society have a strong effect on herd immunity near the homophily value 0.8 and on infection peak near the homophily value 0.7.

**Figure 4:**
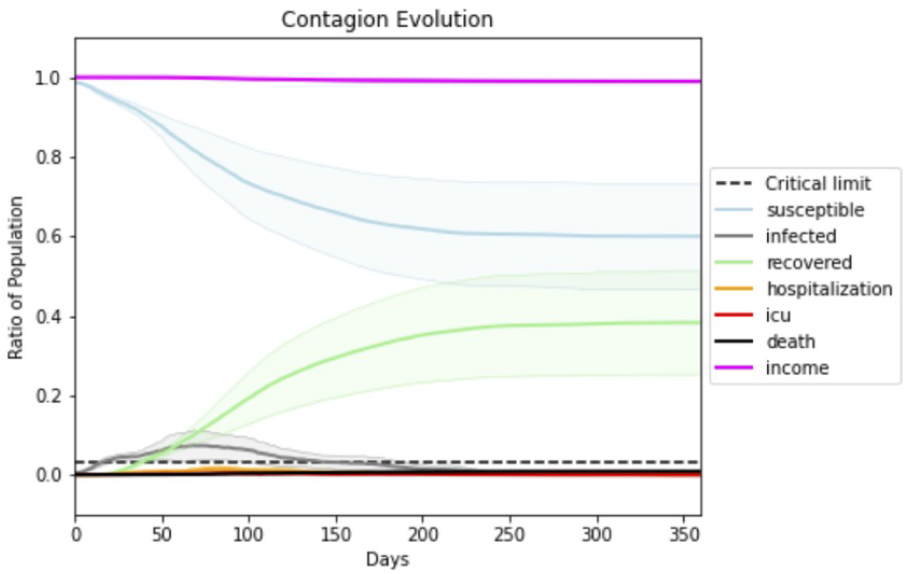
The dark green curve represents the ratio of the population that has recovered from Covid-19. The herd immunity level corresponds to the height at which this curve first begins to flatten and the society becomes resilient against exogenous infections. The dark grey curve represents the current number of infections. All darker-hued curves are based on averaged outputs from 20 runs at the same parameter values, with light-hued envelopes representing 95% confidence intervals. The peak of the grey infection curve and the inflection point of the green curve, which provides an estimate of herd immunity, are the dependent variables of the first study. While herd immunity occurs around 0.2, more of the population are infected as the disease peters out.

**Figure 5:**
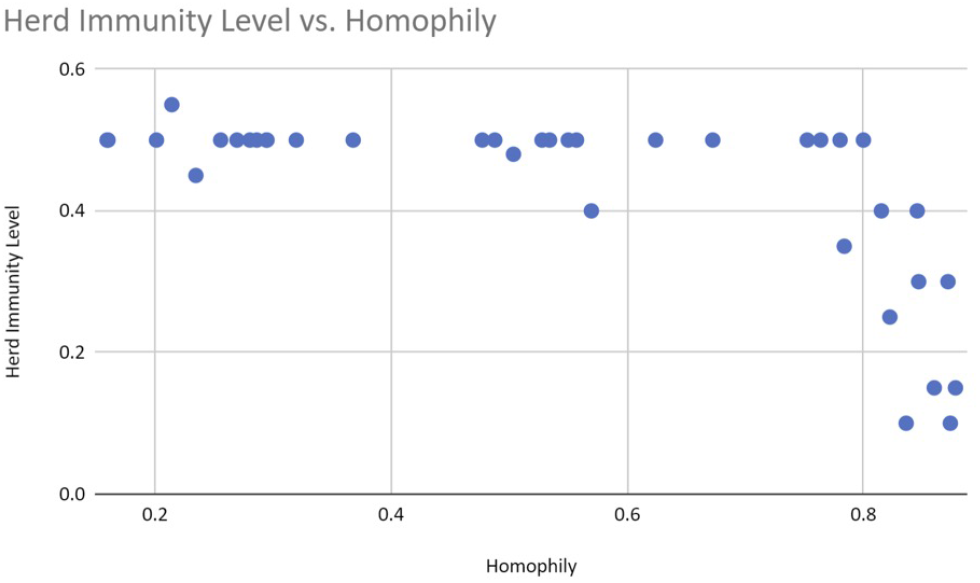
The herd immunity level decreases drastically as homophily levels reach a threshold of 0.8

**Figure 6:**
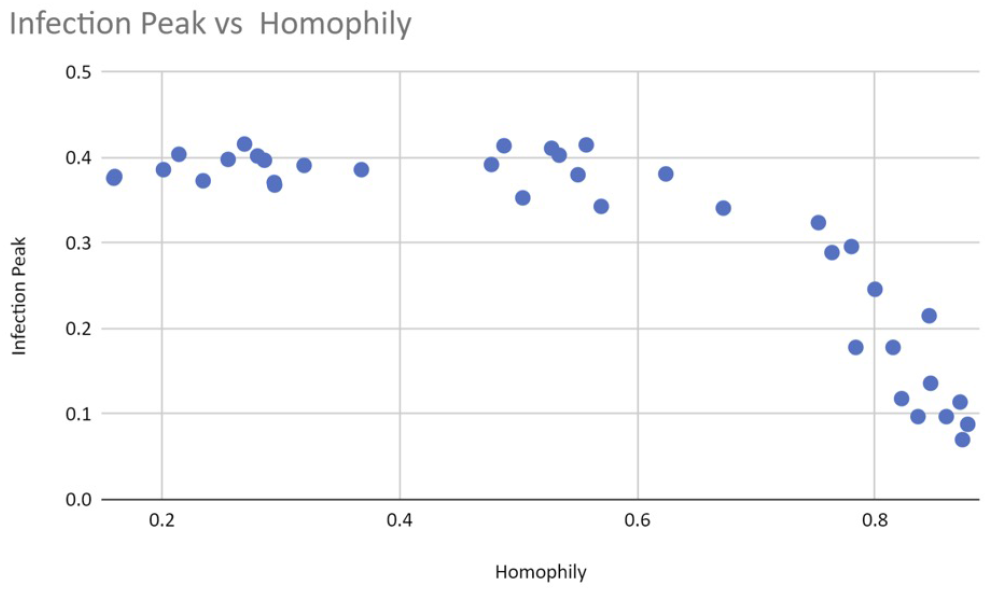
The peak of infection (grey line in time series graphs) decreases drastically as homophily passes the threshold value of 0.7.

We posit that the reason for these threshold effects is initial exponential growth of the largest connected component in the network, called the “giant component” in graph theory. We argue that nonlinear infection rates explode beyond critical thresholds due to the creation of bridges between large, previously isolated sub-components. Creating a bridge between two isolated nodes increases potential infection by one unit; creating a bridge between two isolated node pairs by two units; creating a bridge between two isolated node foursomes, by four units and so on. Thus, exponential increases result from the merging of very large sub-components.

As homophily decreases, connections form between communities that have never been connected before, such that when there are few connections between communities, each new one matters more. Larger groups of people can infect each other with each new connection when communities have few connections to begin with, making the first bridges the most important. Bridging communities is the reason why superspreader events can multiply the effects of diseases on a society wide scale.

As illustrated in Figure 7, rapid growth of the giant component appears to occur simultaneously with a sudden change of infection. This rapid growth also appears to coincide with the functional behavioral thresholds we saw in Figures 5 and 6. Figure 7 displays results with temperatures corresponding to homophily levels spanning the threshold values. The figures display three sets of 20 runs each, each with the same parameters except for very small changes in temperature, and added information about the giant component. The giant component grows more rapidly when there exist sufficient random links between communities due to decreased homophily. The percentage of disconnections also changes, decreasing from ∼25% to ∼0%, as does the herd immunity level, which increases from ∼10% of the population below the threshold value to ∼40% above the value.

**Figure 7:**
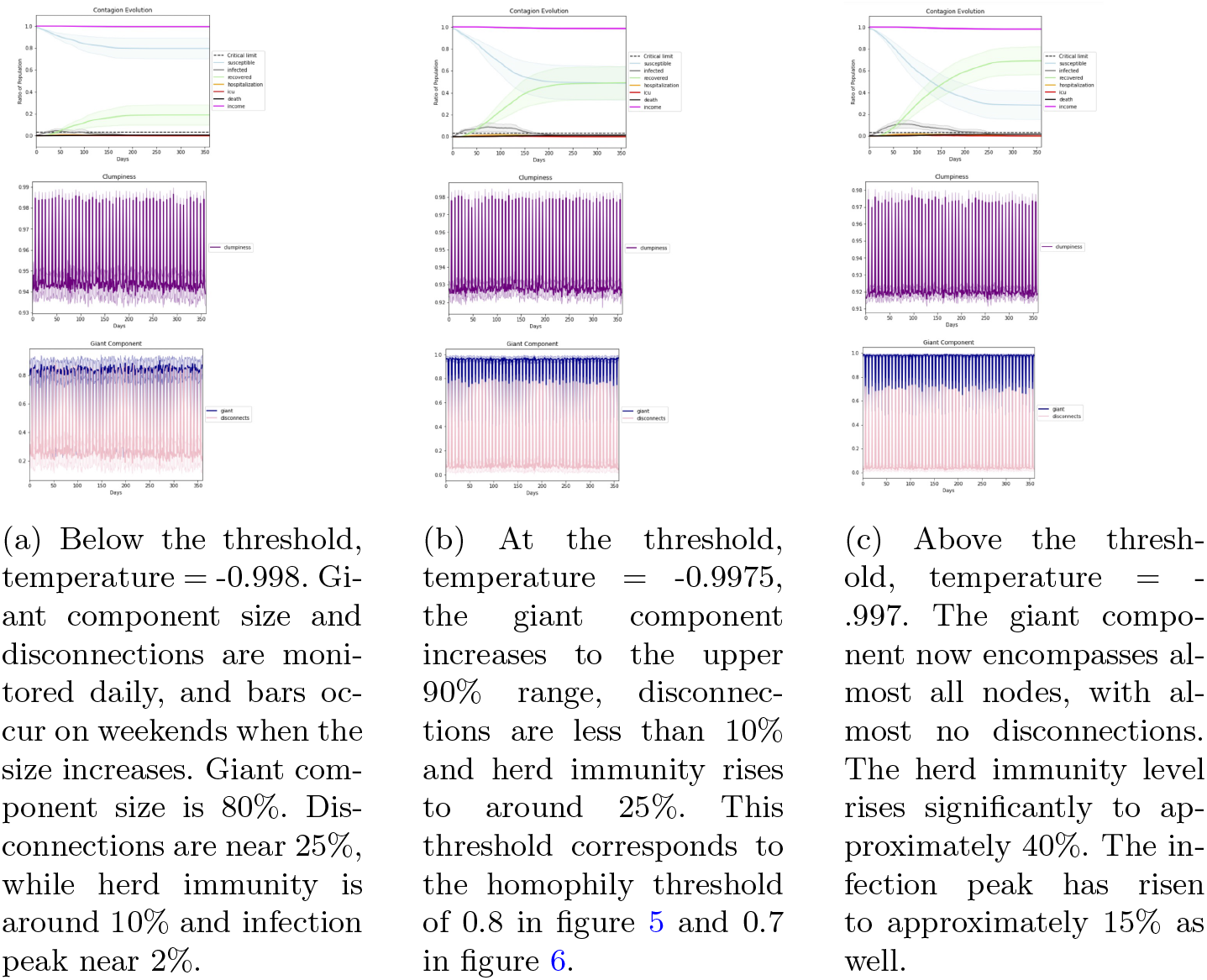
Giant component formation

This result has implications for public health protection strategies. Since clumpiness is calculated from a location graph, it is best to not have labor categories based on demographic characteristics such as race or ethnicity, as it is crucial to protect those individuals who bridge across communities.

A closer look at the data suggests that homophily is nonlinearly correlated with clumpiness (see figure 8), but that clumpiness affects infection and herd immunity levels linearly (see figures 9 and 10). This result makes sense since clumpiness is measured as a number of hops. Formation of the giant component causes this number to increase quickly. The number of hops that separate people is more directly related to infection ability, however, and so should rise only linearly with infection.

**Figure 8:**
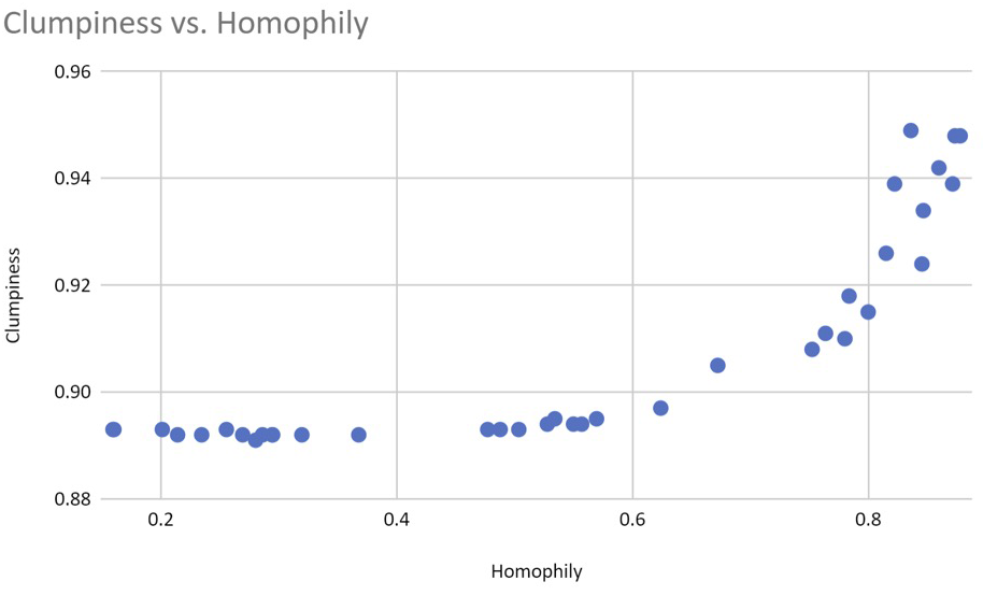
Homophily above the 0.8 similarity level affects clumpiness, non-linearly

**Figure 9:**
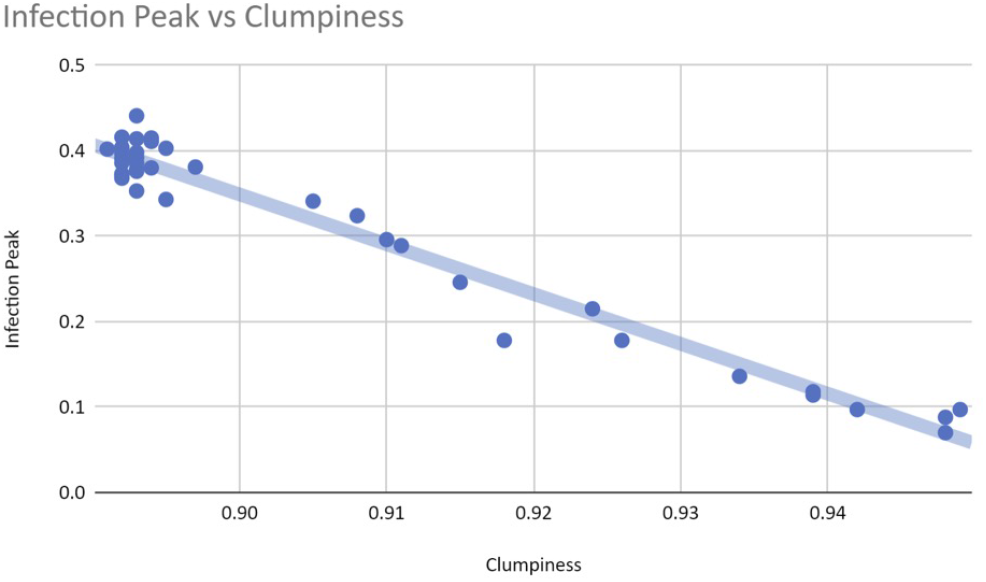
Clumpiness affects the infection peak linearly.

**Figure 10:**
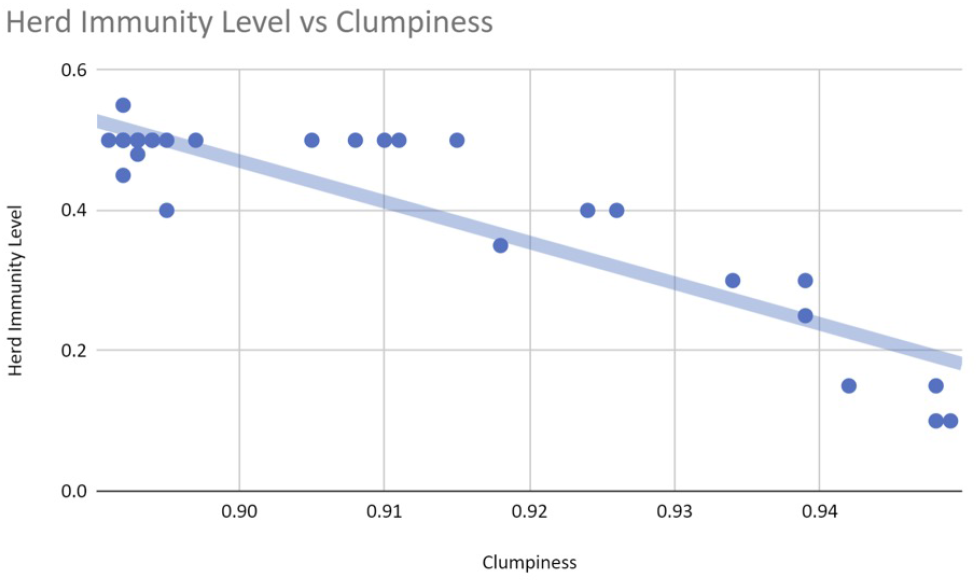
Clumpiness affects the herd immunity levels close to linearly.

The linearity with which clumpiness relates to infection makes it a good monitor and predictor of infection. We can access data on which classes of people typically visit which types of locations, and use this information to infer the level of clumpiness resulting from homophily. Due to the linear relationship between homophily and future infection, we can then predict infection in society. Even in countries with good infection tracking, generalizing specific data into propositions regarding types of persons in types of locations will help with the use of data from specific locations to predict infection in similar locations (via expression in terms of the clumpiness measure). Were we to use this information to create charts similar to Figures 5 and 6, we would know to be patient and continue policies that increase clumpiness near the tipping points. We would know how to time policies for the greatest effect. Figure 11 shows an online clumpiness dashboard, that tells the daily amount of clumpiness, including both from natural homophily, and from policy changes.

**Figure 11:**
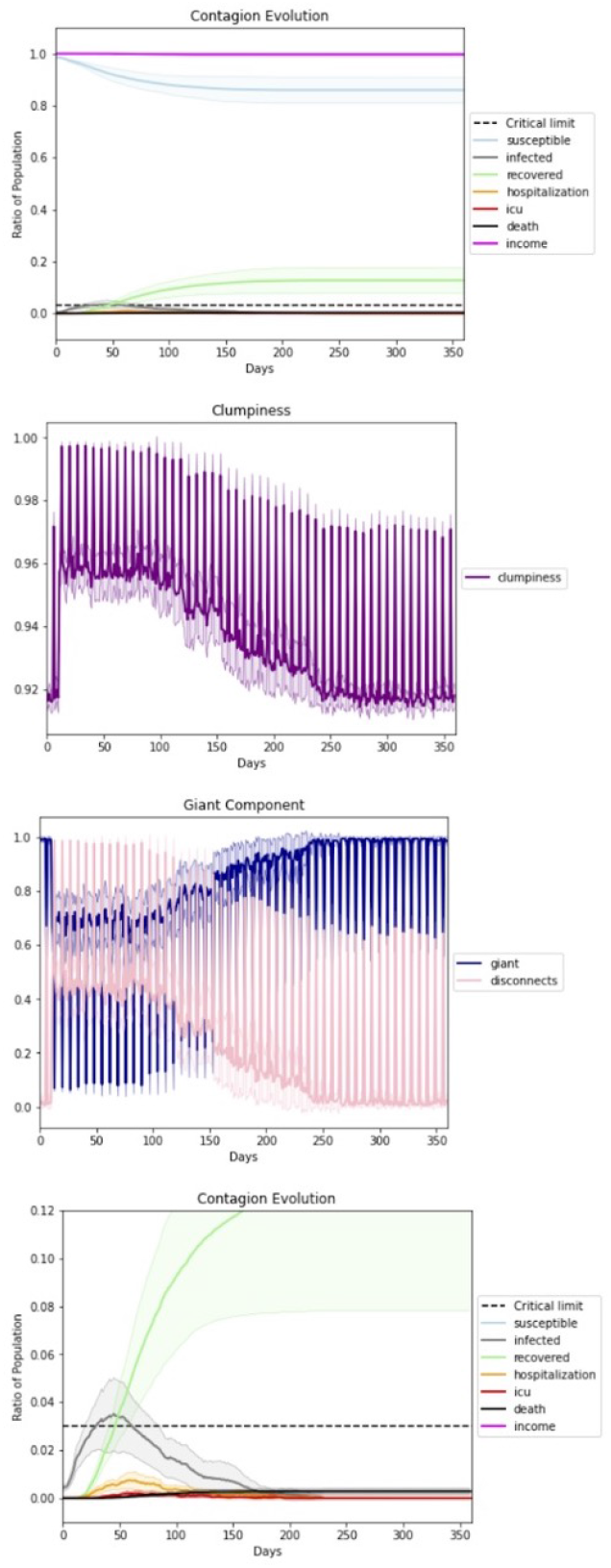
Clumpiness as a policy-maker’s online dashboard for the state of social distancing during a lockdown. Clumpiness, the second graph, monitors the scenario above it, which is zoomed in at the bottom. The spikes in clumpiness represent weekends, when people in our simulation are less liklely to leave their homes. The giant component and the percentage of disconnections, network phenomena that measure clumpiness due to disconnected nodes, are shown. In the twenty runs that these charts illustrate we see a lockdown ending on day 75. The lockdown causes clumpiness to increase. We see clumpiness decrease back to its normal clumpiness value for its homophily level, once the lockdown is removed.

Clumpiness is predictive. This dashboard can provide future forecasts and present maps of predicted future hot spots. The hot spots indicate predictions of where the disease will show up next based on social structure.

### 5.2 Study 2: Clumpiness as a Predictor of Disease Spread

Our second study examines how the individual version of clumpiness, hoprank, can help predict the trajectory of a disease on a fine-grained level. We explore, in particular, whether we can use clumpiness to predict the order in which the disease will arrive at particular schools, workplaces, and restaurants.

Hoprank is a measure of clumpiness on an agent/community-to-agent/community basis. It is calculated by sampling the most probable path between persons, locations, or groups to other non-overlapping persons, locations, or groups. Although hopranking is a sampled and inexact form of contract tracing, involving categories of individuals and locations, it can work without the thorough and detailed data needed for full contract tracing.

We set up our runs so that 20 percent of the communities have active infections. We run tests with random samples of 100 other entities for each hopranked entity, without knowledge of which communities have current infections, and then with infected community samples of 100, 10 and 1 other entities for each hopranked entity. While pagerank requires a complete model of the network, hoprank needs only a number of samples which draw the most probable paths between agents. The 10 and 1 sample runs demonstrate that the measure is robust in conditions in which data availability is minimal. By varying temperatures between −1.0 and −0.99, our study spans homphilly levels from low natural levels of herd immunity to high levels.

In this study we rank all agents in the network on day one with hoprank and, for a baseline comparison, with pagerank. We choose pagerank because it has the advantage of representing the probabilities of all possible packet paths, but the disadvantage of not being able to represent time until arrival. These rankings predict the order in which the disease will arrive at buildings. We let the simulation play out, and see the order in which the disease actually shows up in buildings. Since people tend to habitually attend the same places, their early behavior in the simulation can be used to construct a model of where the disease may travel later. Prediction capability with partial data is important in vaccine prioritization.

We first compute hoprank to random other locations and persons in the simulation, putting it on a level playing field with pagerank, since pagerank does not take into account which communities are infected. Without knowledge of the communities that are infected, hoprank and pagerank perform about the same: Hoprank’s average rank correlation coefficient across all temperatures is 0.38 while Pagerank’s is 0.36. However, in the all the scenarios in which samples are taken from the community where infection is known to occur (and not necessarily of infected individuals in that community) our results demonstrate that hoprank has a considerably stronger positive correlation between the rank of predicted and actual orderings than does Pagerank (see Figure 12 and Table 1).

**Table 1:** Hoprank’s Average Rank Correlation compared to the true order of infection is consistently higher than that of Pagerank when Hoprank’s samples are taken from infected communities, and is equal to Pagerank’s when they are not. The averages are shown across temperatures. T test shows significant results at the p= .01 level when a small sample size of 10 from members of the infected community is taken, indicating that hoprank is robust to small samples of community members whose individual infection state is unknown.

| Sample Size | Sample Origin | Hoprank Correlation | Pagerank Correlation | Pairwise t-Test p-value |
| --- | --- | --- | --- | --- |
| 100 | random | 0.388 | 0.366 | 0.450 |
| 100 | infected community | 0.456 | 0.341 | 0.006 |
| 10 | infected community | 0.498 | 0.307 | 0.010 |
| 1 | infected community | 0.484 | 0.392 | 0.243 |

**Figure 12:**
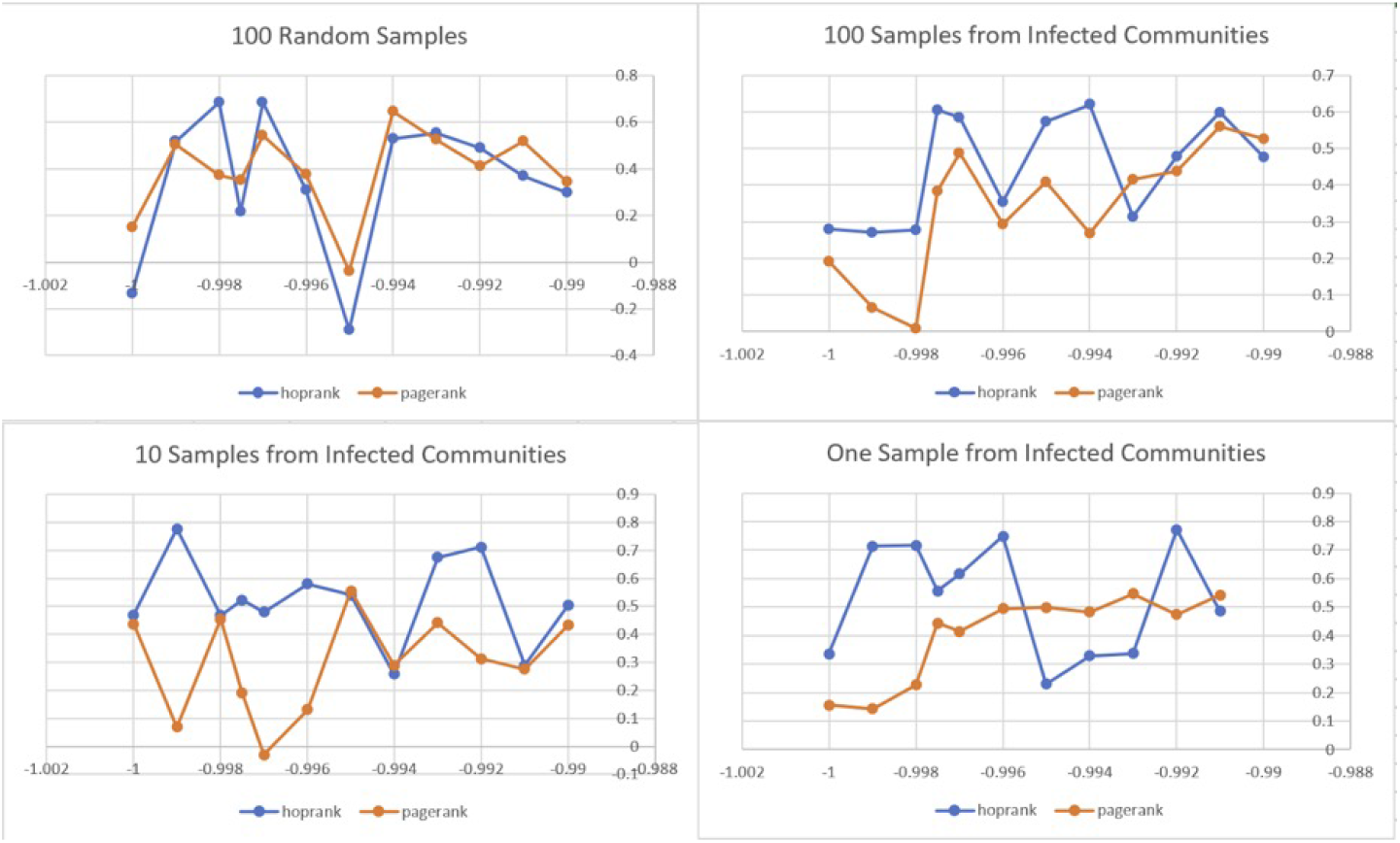
Pairwise comparisons of Hoprank’s and Pagerank’s Rank Correlation Coefficients, at a variety of temperatures and sample sizes. Temperature is on the horizontal axis and Rank Correlation Coefficient is on the vertical axis. Hoprank performs better than pagerank when hoprank’s samples are taken from the infected community. While runs are unique (for example, infection dies out earlier in some than others) each individual pairwise comparison is significant at an *α* = 0.99 level.

**Figure 13:**
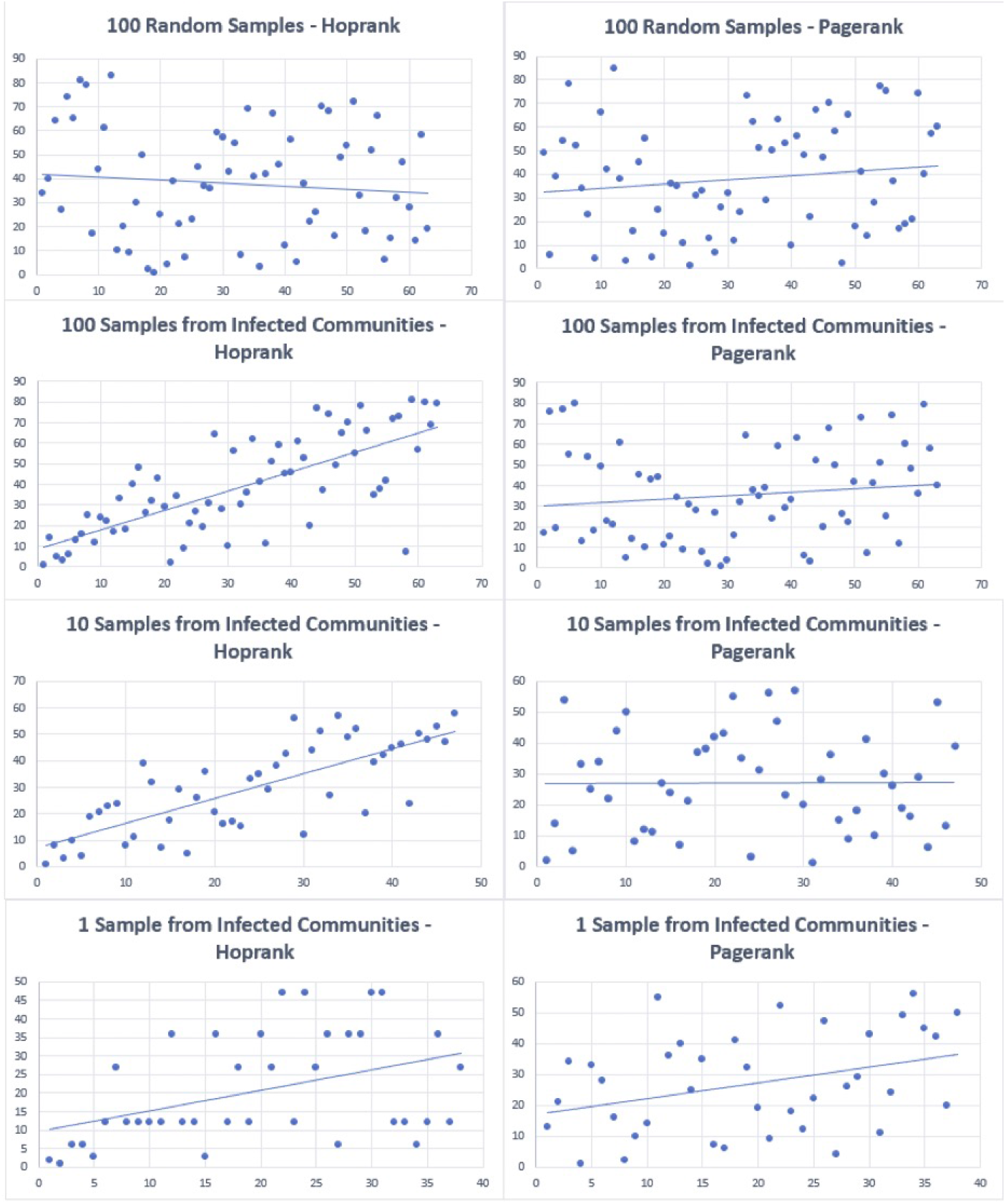
Four runs comparing the Hoprank and Pagerank algorithms using the data for Figure 12 corresponding to temperature −0.9975 and to multiple sampling scenarios. Each dot represents a classroom, office or restaurant, with the actual orders of infection occurrence along the horizontal axis, and predictions, by both algorithms, of the orders of infection occurrence along the vertical axis. With 100 random samples the rank correlation for hoprank is 0.21 and for pagerank is 0.35. With 100 samples from the infected community the rank correlation is 0.60 for hoprank and 0.38 for pagerank. In the 10 sample scenario, the rank correlation coefficient is 0.52 for hoprank and 0.18 for pagerank, and in the 1 sample scenario, the rank correlation coefficient is 0.55 for hoprank and 0.44 for pagerank.

### 5.3 Study 3: Determine the effect of Homophily Behavioral Networks on herd immunity level and on infection

In this study we move from the physical proximity network to a network of ideas. The similarity of agents and their friends depends on the scenario temperature, and the number of those friends depends on the agents’ personal extroversion parameters. Agents with a tendency to herd follow their friends’ risk tolerance levels more than their own. Having a high risk tolerance might lead them to break lockdowns and go to work or to bars and restaurants.

To study the relationship between herding and risk tolerance on compliance, we test, for the agents of a scenario, high and low tendencies to herd against high and low risk tolerance in a lockdown environment. We generate 20 runs of each combination and at ten different temperatures. We find that herding has a strong and significant protective effect in high risk scenarios under lockdown when looking at all temperatures, with an effect size of 130% on the herd immunity threshold, significant at the 0.000008 p level in a paired t test, and described as a large effect size by Cohen’s equation. Table 2 shows the data by temperature for herd immunity and table 3 shows the data by temperature for infection peak. It also has an effect size of 122% on the infection peak, also significant at the 0.025 p level in a paired t test. In both cases, the effect appears to be stronger at hotter temperatures, indicating that having friends that are different from you allows the protective effect to spread to other communities.

**Table 2:** At eight out of ten temperatures, low herding has a higher herd immunity threshold, which we use the recovered peak statistic to represent. Its effect size is over 130%, significant at the p=0.000008 level, in a paired *t* test.

| Temperature | Herd Immunity<br>High Herd | Herd Immunity<br>Low Herd | Effect Size |
| --- | --- | --- | --- |
| -1 | 0.015 | 0.020 | 1.33 |
| -0.999 | 0.16 | 0.020 | 1.25 |
| -0.998 | 0.017 | 0.020 | 1.18 |
| -0.9975 | 0.018 | 0.025 | 1.39 |
| -0.997 | 0.020 | 0.030 | 1.50 |
| -0.996 | 0.017 | 0.025 | 1.47 |
| -0.995 | 0.028 | 0.035 | 1.25 |
| -0.994 | 0.025 | 0.030 | 1.20 |
| -0.993 | 0.025 | 0.030 | 1.20 |
| -0.992 | 0.020 | 0.025 | 1.25 |
| **mean** | 0.020 | 0.026 | **1.30** |
| **stdev** | 0.004 | 0.005 | 0.12 |

**Table 3:** Infected peak is greater in low herd than high herd scenarios, for eight out of ten temperature values, with an effect size of 122% significant at the p= 0.025 level in a paired t test.

| Temperature | Infected Peak<br>High Herd | Infected Peak<br>Low Herd | Effect Size |
| --- | --- | --- | --- |
| -1 | 0.0210 | 0.0230 | 1.10 |
| -0.999 | 0.0280 | 0.0280 | 1.00 |
| -0.998 | 0.0230 | 0.0290 | 1.26 |
| -0.9975 | 0.0280 | 0.0320 | 1.14 |
| -0.997 | 0.0250 | 0.0290 | 1.16 |
| -0.996 | 0.0210 | 0.0330 | 1.57 |
| -0.995 | 0.0270 | 0.0250 | 0.93 |
| -0.994 | 0.0260 | 0.370 | 1.42 |
| -0.993 | 0.0270 | 0.0260 | 0.96 |
| -0.992 | 0.0270 | 0.0450 | 1.67 |
| **mean** | 0.0253 | 0.0307 | **1.22** |
| **stdev** | 0.0027 | 0.0065 | 0.26 |

Somewhat counterintuitively, communities that are low risk in a high risk population are able to protect themselves by copying each other, but high risk individuals do not recruit low risk relations into adopting high risk behavior in equal measure. There was no significant difference between groups in the lower risk scenarios, meaning that low risk individuals can only recruit high risk individuals if they are in the majority. Perhaps the fact that in order to be copied agents must have recent behavioral histories, and high risk individuals take themselves out of commission by getting sick at higher rates than do low risk individuals. We should expect a similar phenomenon in the real world where infections run rampant, as they do at high temperatures in the simulated world.

## 6 Conclusion and Future Work

We introduce a simulation to demonstrate the usefulness of a novel metric, clumpiness, and offer results that show predictive power of social-behavioral variables on epidemiological ones. The results also suggest the importance of bridges that make a case for changing the infection state directly through changing homophily by protecting bridges between communities. We further show how to use clumpiness to indirectly change the state by using it to predict and adjust vaccination prioritization. We create a dashboard that can help policymakers by alerting them to social distancing states and projecting the disease trajectory. We also look at natural protection afforded by homophilic communities where memetic behavior is concerned. Such knowledge is necessary for effective messaging campaigns.

Importantly, this measure of clumpiness is a general social network measure that describes the frictive surface upon which packets may travel, be they viruses in a physical proximity network or memes in a proximity social network. Looking at the speed of packet reproduction along these networks is a framework of analysis for problematic polarizing social networks as well as a potential framework for formulation of solution.

One possible direction for future research is to use intelligent agents to model epidemiological messaging. We would give agents a “mind” with features that are important to modelling sense making, such as cognitive dissonance and reinforcement learning. By enabling more realistic models of belief and behavior dissemination through networks, both naturally and in the presence of information operation campaigns, the use of such agents would allow us to model how information operation campaigns convince and incentivize victims.

Such information would allow us to tackle one of the most important issues affecting epidemiological public policy: adversarial disinformation campaigns. By accessing real world data on social media platforms, we could match detailed profiles with locations. In the current study, we describe a general highway upon which packets may travel and demonstrate that this highway is useful for predicting where and when a virus may travel. Similar highways could help predict paths for idea transmission and map the spread of divisive disinformation.

For example, adversaries may exploit the information highway by causing small increases in homophily that result in large changes in transmission, creating information bubbles that isolate ideas into different groups, so that there is a different reality for each group. If public policy makers were to see the information highways of social media in this light, they might be able to better regulate social media companies to protect the public against information warfare.

## Data Availability

This was a simulation. Parameters were set based upon data published in openly available sources. All other data was internally generated.

## Appendices

### A Demographic Stratification

It is now clear that COVID-19 strikes particular demographic strata much harder than others. To better model this phenomenon, we have explicitly incorporated the following key factors in the initial implementation of our model:

- Multiple classes of individuals:
  – Infants and toddlers
  – K-12 school aged children
  – Working-age adults
  – Elders
- Sub-classes for working-age adults:
  – Office workers (have a work location but can work from home)
  – House-bound individuals (whether they work from home, are trophy househusbands, etc. seems not to matter much)
  – Factory workers (can’t work from home but don’t meet non-coworkers during their work hours)
  – Retail workers (can’t work from home AND meet non-coworkers during their work hours)
  – Essential workers
  – Teachers
  – Hospital workers
- Location classes including:
  – Houses;
  – Apartments in a building;
  – Offices in a building;
  – Schools;
  – Hospitals (along with hospital system capacity)
  – Factories
  – And multiple restaurant types
    * Fast food restaurants;
    * Full-service restaurants;
    * Bars.
- Economic impact modeling that factors in the above classes and sub-classes.

We have also incorporated into our models a reasonable diversity of relatively simple personality characteristics of the simulated agents. While assuming that everyone will follow social distancing guidelines isn’t realistic so is randomly deciding who doesn’t. To obtain expected individual behaviors, we endow each simulated individual with an extensible set of personality parameters including:

- A risk tolerance parameter;
- And a “herding behavior” parameter that makes agents more or less likely to adopt given behavior(s) (in compliance or defiance of official policies) based on the behavior of their peers.

#### A.1 A Flexible and Extensible Framework

We designed our simulation framework to be flexible and extensible. Most notably, we architectured our system with the introduction of intelligent and adaptive agents in mind. When the time comes, the various personality parameters will form key inputs to each learning agent’s utility function.

Future versions will also include individual health attributes – most simply modeled via an overall health parameter, which can be reduced by the presence of comorbidities and immuno-suppressed individuals and increased for fit individuals.

Our modular architecture will further allow for easy and quick implementation of periodically-updated region-specific demographic data, and inclusion of additional locations such as:

- Retail shops;
- Grocery Stores;
- Hospitals (along with hospital system capacity);
- Factories;
- And recreational gathering spots.

### B Parameters of Interest

#### B.1 Disease Parameters

Prior to setting our agent-based disease parameters, we found the most current COVID-19 data we could. Most of this data, for example for *R*_0_, pertained directly to macro-scale modeling and hence could not be used directly within our agent-based approach. To handle this situation, we adjusted agent-level parameters so that such macro-scale social-level parameter values emerged naturally.

We display disease and hospital capacity parameter settings in tables 4 and 5.

**Table 4:** Essential Pandemic Default Parameters

| Parameter | Distribution |
| --- | --- |
| Latency Period | Gamma( Shape = 3, Scale = 1) |
| Incubation Period | Gamma( Shape = 6, Scale = 1) |
| Mild Period Duration | Gamma( Shape = 14, Scale = 1) |
| Hospitalization Period Duration | Gamma( Shape = 14, Scale = 1) |
| Spreading Rate | Normal( Mean = 3.155, Std Dev = 1.7) |

**Table 5:** Disease and Medical Facility Probabilities

| Parameter | Probability |
| --- | --- |
| Initial Infection Rate | 0.01 |
| Immune Rate | 0.01 |
| Hospitalization Capacity | 0.05 |
| ICU Capacity | 0.03 |
| Symptomatic Isolation Rate | 0.0 |
| Asymptomatic Contagion Probability | 0.1 |

#### B.2 Distancing and Behavioral Parameters

Our model also includes a variety of parameters to control for individual agent behaviors such as risk tolerance and herding behavior; and for government policy control measures including partial lock-downs and building occupancy measures. We display default parameter settings for these groups of parameters in tables 6 and 7.

**Table 6:** Building Occupancy Parameters

| Parameter | Value |
| --- | --- |
| Allowed Restaurant Capacity | 0.25, 0.5, or 0.75 of normal occupancy |

**Table 7:** Behavioral Parameters

| Parameter | Probability or Distribution |
| --- | --- |
| Mask User Rate | 0.9 |
| Mask Efficacy | 0.6 |
| Risk Tolerance | Beta(Mean = 0.7, Std. Dev = 0.2) |
| Herding Behavior | Beta(Mean = 0.7, Std. Dev = 0.2) |

**Table 8:** Restaurant Contagion Levels

| Percent Occupancy Level | Distancing | Mean $R_0$ | $R_0$ 95% CI |
| --- | --- | --- | --- |
| 25% | $\sim 17$ feet | 0.21 | (0.08, 0.5) |
| 50% | $\sim 6$ feet | 0.3 | (0.11, 0.74) |
| 75% | $\sim 4$ feet | 0.5 | (0.19, 1.23) |

Our model also allows for different contagion probabilities, depending upon multiple factors including building and business locations and types (e.g. offices, bars, sit-down restaurants, fast-food restaurants;) and social distancing effects of imposed occupancy limits (e.g. 25%, 50%, 75% of normal limits.) Finding specific values in the literature for the macro-level reproductive number *R*_0_ proved challenging, partially due its evolving nature as individuals responded to the pandemic. Two meta-analyses [11, 3] provide mean values of 3.15 and 3.32 and 95% CIs of (2.41, 3.90) and (2.81, 3.82) respectively. Applying the effects of physical distancing described in Chu, et al., [8] we obtain rough values for the reproductive number at a distance of one-meter as follows: mean 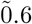; 95% (actually 97.5%) CI ∼ [0.22, 1.48]. We also obtain values at a distance of two meters having mean ∼0.3, and a ∼95%CI of ~ [0.11, 0.74].

Investigating US state reopening rules for restaurants, many states (e.g. Colorado) have rules allowing for the lower of 50% occupancy levels or occupancy level to support a minimum distance between parties of 6 feet. With this in mind, a 50% occupancy level is being used as a proxy for maintenance of 6-foot social distancing. Calculating distancing corresponding to 25% and 100% occupancy involves simple multiplication or division, respectively, of the 6-foot distance by Sqrt(2) 1.414. Applying the rules in the Lancet physical distancing article then gives the following table of rough estimates:

### C The Role of Propaganda

A network *G* exhibiting a high degree of clumpiness can be viewed as consisting of separate subgraphs *S*_*i*_ connected by bridge(s) as illustrated in figures 14 and 15.

**Figure 14:**
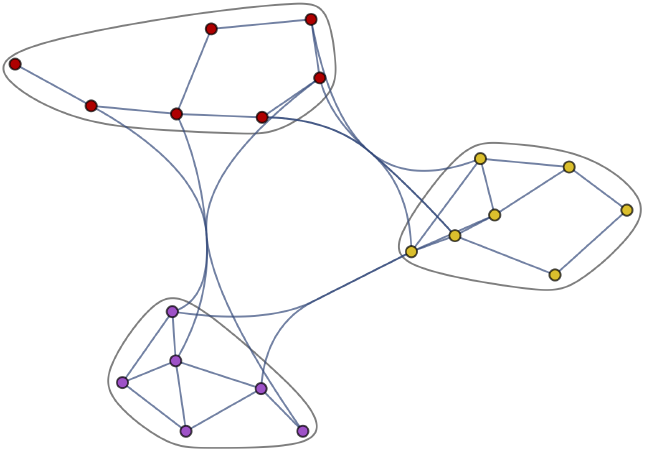
Social Subgraphs

**Figure 15:**
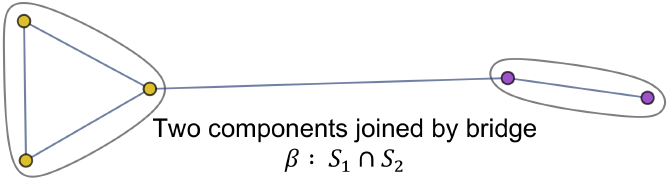
Bridge *β*_12_ Connecting subgraphs *S*_1_ and *S*_2_

**Figure 16:**
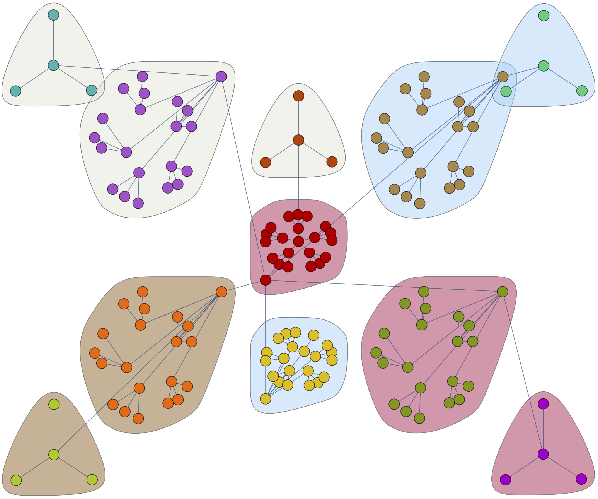
Contagion Connections

**Figure 17:**
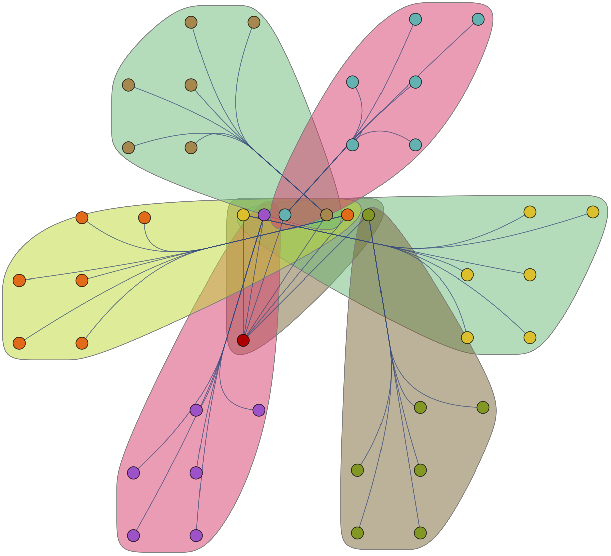
Superspreads

Contagion spreads may be disconnectedly joined by bridges, such that *β*_12_ : *S*_1_ ∩ *S*_2_, *β*_34_ : *S*_3_ ∩ *S*_4_, etc.

However, the most acute peril is posed by superspreads:

Each community *S*_*i*_ exhibits one or more mimetic behavioral practices, which may be motivated by homophily and/or polarization. Behavior as such determines the emergent percolative properties of the spread with respect to some critical threshold *π*_*c*_. Contending mimetic influences may produce within a spread a mixture distribution 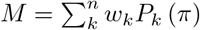.

We have instituted a Propaganda class in our simulation code to study the effects of such mimetic behaviors that more strongly manifest themselves via high risk, high herding individuals.

## Footnotes

1 In more complex studies, clumpiness could occur in conjunction with diversity, but for this first simplistic study, communities consist of colocated individuals who share similar social demographic traits.

## Notes

### Competing Interest Statement

The authors have declared no competing interest.

### Funding Statement

All authors except for James Boyd were paid for their work by their employer SingularityNET Foundation. No external funding was received.

### Author Declarations

No human subjects were involved in this research.

